# Long term use and replacement of near vision glasses for presbyopia, in Punjab, Pakistan: a cross-sectional survey

**DOI:** 10.64898/2026.08.15.26360501

**Authors:** Muhammad Moin, Rizwan Younas, Shahid Maqbool, Zahid Hussain Awan, Muhammad Bilal, Marzieh Katibeh, Elanor Watts, Sergio Latorre-Arteaga, Patricia Elaine Freels, Abigail Steinberg, Andrew Bastawrous

## Abstract

**Background:** An estimated 826 million people have avoidable near vision impairment (NVI) due to lack of access to near vision glasses for presbyopia. Glasses use, uptake and replacement is essential to sustained NVI correction. Here we explore the uptake of second and subsequent pairs of near vision glasses and the willingness to pay.

**Methods:** In this cross-sectional survey, 276 individuals who received near vision glasses through eye health screening programs in Punjab province, Pakistan from 2020 to 2022, were surveyed between February and July 2025, i.e. 3–5 years later.

**Results:** 92.0% (n=254) of respondents were still using near vision glasses: a purchased replacement (41.3%, n=114), a free replacement (30.8%, n=85), or the original pair (19.9%, n=55). Male gender, higher age, and personal income were strongly associated with purchasing additional pairs, while women and those economically dependent on others relied more on free provision (all p values <0.05). Accessibility, indicated by shorter travel times and awareness of supply sources, also played a significant role in sustained replacement. Over 98% of participants expected to obtain (purchase or receive) a new pair of glasses in the future, and over 94% of participants expected to purchase their next pair. The mean willingness-to-pay (WTP) was 353 PKR (95% CI: 327–379) // 1.25 USD (95% CI: 1.16-1.34), while the average reported price paid was 416.67 PKR // 1.47 USD (95% CI: 351.39-481.94)).

**Conclusion:** Sustained use of near vision glasses remained high 3-5 years after initial provision. Many recipients had purchased replacements, and willingness and ability to pay were high among participants, suggesting potential for sustained demand beyond initial subsidized distribution.

## Background

Presbyopia is the gradual loss of near vision with age, which, if left uncorrected, causes near vision impairment (NVI), defined as near visual acuity worse than N6 at 40 cm.^1^ It is the most common cause of visual impairment worldwide, affecting an estimated 826^3^ million people, a number expected to continue to rise due to population aging and growth. The annual global productivity loss attributable to uncorrected presbyopia is estimated at US$25.4 billion.^4,5^ Beyond economic costs and livelihoods, the condition has substantial adverse effects on individuals’ wellbeing, independence and quality of life.^6,7^

In most cases, presbyopia can be effectively corrected with near vision glasses.^8^ Ready-made readers, which provide identical correction in both eyes, are suitable for the majority of affected individuals without other ocular conditions or significant distance refractive error. These can be provided quickly and inexpensively, with or without healthcare worker support. Nevertheless, nearly half of people worldwide who need near vision glasses lack access,^3^ owing to factors such as geographic isolation, limited service availability or affordability, and lack of awareness among both the public and healthcare providers. Within health systems, interventions for presbyopia have often been overlooked or deprioritized, sometimes due to concerns about quality or the perceived impact on other eye care services.

In a recent retrospective analysis of 1,069,372 individuals aged 35+ who attended near vision screening in Peek-powered programs, across nine countries from Jan 2022–Aug 2024, those using near vision glasses were significantly more likely to have healthy eyes or to meet their eye care needs: a subset of this data has been previously published.^9^ Specifically, 88% required no further referral or successfully attended referrals when needed, compared with 74% among those without glasses. Uncorrected presbyopia emerged as the most common eye health problem in adults over 50 years of age. These findings suggest that provision of reading glasses at community and primary levels not only improves near vision but may also facilitate greater engagement with eye care services, while also helping specialized services focus on more complex, sight-threatening conditions such as cataracts. This model is in line with the WHO Competency-based Refractive Error Teams (CRET) framework^8^ which encourages provision of presbyopia correction at the community level.

The sustained use, uptake, and replacement of near vision glasses are critical to maintaining these benefits, yet these factors remain largely unstudied. To address this gap, the present study investigates the uptake of second and subsequent pairs of near vision glasses and explores the barriers, satisfaction, and willingness to pay for additional pairs among individuals who received near vision glasses through CBM Peek-powered programs in Punjab province, Pakistan, three to five years previously. By examining the factors influencing long-term use and access, this research aims to identify strategies to enhance the availability and consistent uptake of near vision glasses in these communities, and to demonstrate whether a concerted push to provide people with their first pair, could lead to self-sustaining uptake thereafter.

## Methods

### Study Population and Design

Between February and July 2025, a cross-sectional door-to-door or telephone survey was conducted, with a sample of 276 participants selected from 943 individuals who had received free or subsidized near vision glasses between 2020 and 2022 through Community Eye Health programs in Punjab province.

### Eligibility Criteria

- Participant of CBM Peek-powered Community Eye Health programs in Punjab province, Pakistan, between 2020 and 2022
- Recipient of free or subsidized near vision glasses
- Consented to participate

### Data Collection and Management

Data from the telephone survey was collected by trained personnel from the College of Ophthalmology and Allied Vision Sciences (COAVS) in Punjab. Standardized questionnaires assessed demographics, employment, income, current glasses use and replacement history, self-perceived barriers to replacement, motivators, replacement sources and costs, strength adjustments, vision perception, and willingness to pay. The survey questions are available in Supplementary Material.

Data was exported in csv and MS-Excel (Microsoft Corp., WA United States) format. Data was recorded digitally within password-protected devices, and will be stored for a minimum of 5 years after the completion of the study, including the follow-up period.

### Data Analysis

Data were analyzed using Stata/IC 14.2 (StataCorps LLC, TX United States). Participants were categorized in 4 groups based on replacement and current use:

1a. Still using the original pair

1b. Using new glasses (purchased)

1c. Using new glasses (received for free)

Subtotal 1. Currently using near vision glasses (1a + 1b + 1c)

2. No longer using glasses

Individuals aged under 40 years, as well as those presenting with other eye conditions (including distance refractive error), were not eligible to participate in the survey.

Differences in responses to key survey questions were examined by demographic variables using the chi-squared test.

For multivariate analysis, binary and numeric outcomes were considered. Logistic regression analysis was conducted to explore the independent effects of multiple variables simultaneously. For both chi-squared and logistic regression models, statistical significance was set at p < 0.05.

## Results

### Participant demographics and income

A total of 276 participants with a mean age of 53.2 years (SD: 8.7, range 40-90) were included in the survey. The baseline demographics and income status are presented in Table 1.

**Table 1:** Demographic and economic status among study participants (N=276).

|  |  |  |  |
| --- | --- | --- | --- |
|  |  | Freq. | Percent |
| Gender | Female | 86 | 31.3 |
|  | Male | 189 | 68.7 |
|  | Declined to answer | 1 | 0.4 |
| Age groups | 40-49 | 98 | 35.5 |
|  | 50-59 | 114 | 41.3 |
|  | 60-69 | 52 | 18.8 |
|  | ≥70 | 12 | 4.4 |
| Main source of income | Employed or have income | 174 | 63.0 |
|  | Casual work | 34 | 12.3 |
|  | Farming | 21 | 7.6 |
|  | Pension | 27 | 9.8 |
|  | Regular employment | 35 | 12.7 |
|  | Rental income | 4 | 1.5 |
|  | Self-employment | 34 | 12.3 |
|  | Other | 19 | 6.9 |
|  | Dependent on other | 102 | 37.0 |
| Income (Pakistani Rupees) | All respondents (including people with 0 income) | 231 | 83.7 |
|  | Mean (95% CI) | 21,893 (95% CI: 18,753, 25,033) |  |
|  | Median (P25, P75) | 20,000 (0, 40,000) |  |
|  | Only Respondents with income | 124 | 44.9 |
|  | Mean (95% CI) | 40,785 (95% CI: 37,602, 43,967) |  |
|  | Median (P25, P75) | 39,700 (30,000, 53,000) |  |
| P: percentile |  |  |  |
| *1 Pakistani Rupee equals 0.0035 United States Dollar, per exchange rates correct as of September 28 2025 |  |  |  |

Regarding sources of income, approximately one-third of participants (37.0%) reported being financially dependent on others, while 63.0% of participants (95% CI: 57.1–68.8%) reported being employed or having an income. The mean reported monthly income among all respondents was 21,893 PKR (95% CI: 18,753–25,033) // 77.3 USD (95% CI: 66.2-88.4).

Among 86 female participants, 85 (98.9%) were economically dependent on others, and only one (1.2%) was employed. In contrast, among 188 male participants, 17 (9.0%) were economically dependent, while 171 (91.0%) were employed or had an income.

### Near vision glasses ownership, use and replacement behaviour

Most participants (95.7%; 95% CI: 92.5–97.7%) received their first ever pair of reading glasses through the program. Of the 276 participants, 92.1% (95% CI: 88.2 - 94.9%) currently owned a pair of near vision glasses. This included 41.3% (95% CI: 35.4-47.0%) who had purchased new glasses, 30.8% (95% CI: 25.7-36.5%) who had new glasses which were obtained for free, and 19.9% (95% CI: 15.7-25.2%) who were still using their original pair. Only a small percentage (7.9%, (95% CI: 5.3% to 11.9%)) reported they had stopped using near vision glasses. Approximately one-third (31.5%; 95% CI: 26.1–37.4%) still owned the glasses obtained from the program, though some of them had also acquired new glasses. In this analysis, such cases were classified under the “second pair” category (new–purchased or new–free).

Figure 1 illustrates the current distribution of near vision glasses used/obtained among study participants, the main use of near vision glasses among participants, with reading as the most commonly reported main use (33.3%), and the frequency of use.

**Figure 1:**
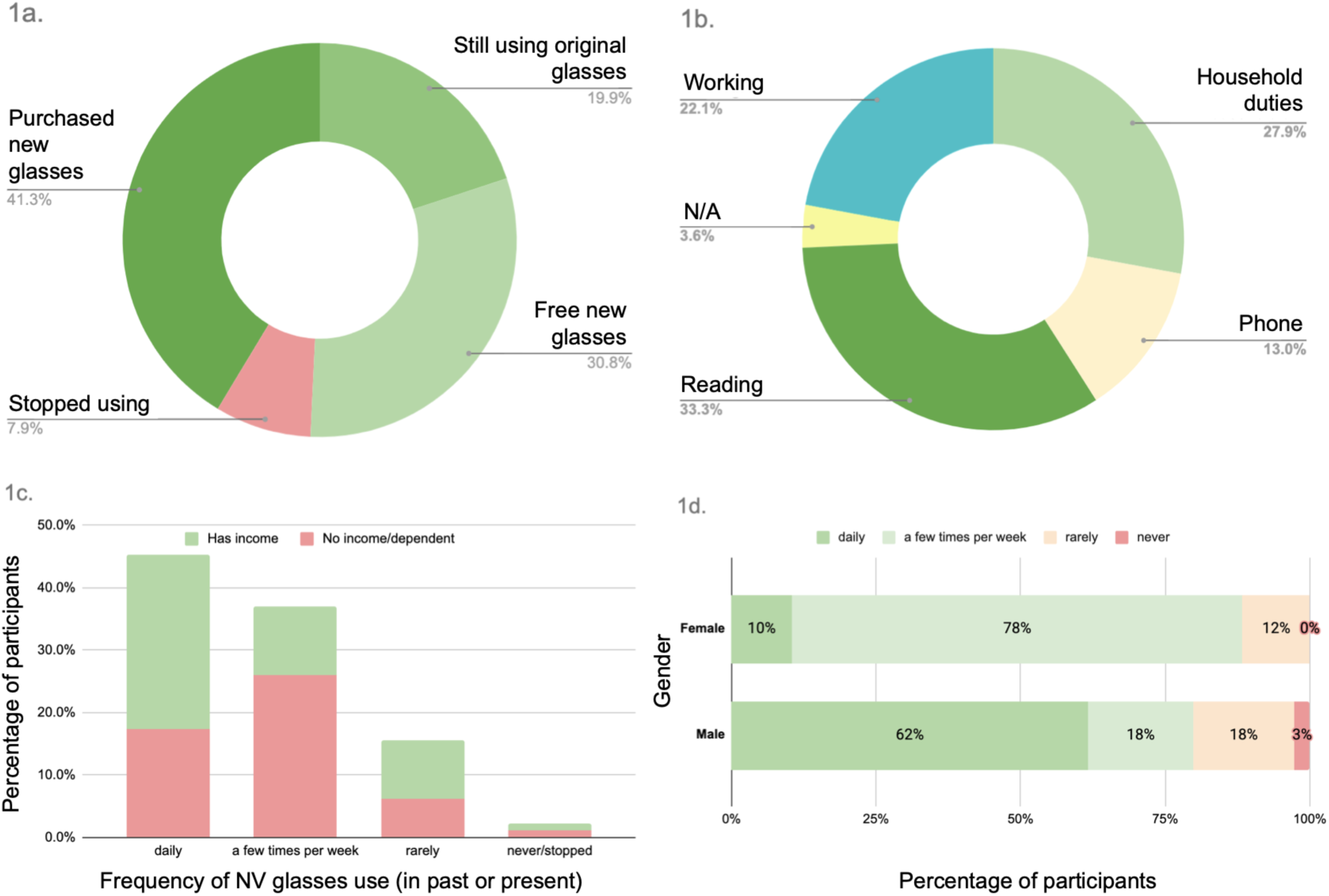
a) Current near vision glasses ownership and replacement status, b) Most common use of near vision glasses, c) Frequency of use of near vision glasses, by income status, d) Frequency of use of near vision glasses, by gender.

The vast majority of participants (97.8%) reported a need for near vision glasses use.

Overall, individuals with income were more likely to use reading glasses daily, while those without income are more likely to use them less frequently (a few times per week). 272 (92.0%) were still wearing near vision glasses, either first of subsequent pairs. Discontinuation of use of the first pair was due to barriers including damage (70.0%), loss (15.3%), discomfort/disliking the glasses (6.6%) or others taking the glasses (2.7%). However, 68.7% (n=189/272) of people who were no longer wearing their first pair 49.2% (n=93) have gone on to purchase and use subsequent pairs.

Since the program, nearly three-quarters (72.1%, 95% CI: 66.7%–77.4%) obtained another pair of near vision glasses: 43.7% (n=85) received them for free, while 57.3% (n=114) purchased them themselves.

Figure 2 shows the status of near vision glasses ownership 3-5 years after receiving free near vision glasses through eye health screening programs in Pakistan in 2020-2022.

**Figure 2:**
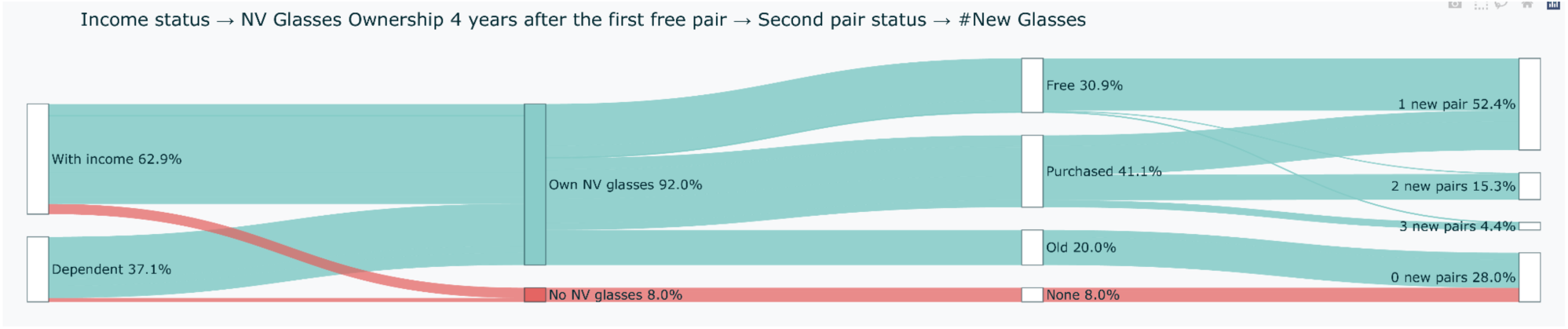
Participant income status, subsequent near vision glasses use 3-5 years after program provision, and replacement status, including number of replacement pairs.

### Future expectations and willingness to pay

Over 98% of participants are expecting to obtain (purchase or receive) a new pair of glasses in the future, and over 94% of participants expected to purchase their next pair.

When asked about prices, participants expected the price of new near vision glasses to be 504 PKR on average (95% CI: 449–559) // 1.79 USD (95% CI: 1.59-1.98). The mean willingness-to-pay (WTP) was 353 PKR (95% CI: 327–379) // 1.25 USD (95% CI: 1.16-1.34).

Among those who did pay for a new pair, the average reported price paid was 416.67 PKR // 1.47 USD (95% CI: 351.39-481.94)).

### Comparison across groups - univariate analysis

Results of univariate analysis are summarized in Table 2. Men were more likely to purchase new glasses (54.0%) and also more likely to have stopped using glasses (11.1%), compared to women, who predominantly received free glasses (74.4%) and had low rates of stopping use (1.2%) or purchasing replacements (14.0%).

**Table 2:** Key Outcomes by demographics, economic factors and glasses use.

| Table 2: Key Outcomes by demographics, economic factors and glasses use |  |  |  |  |  |  |
| --- | --- | --- | --- | --- | --- | --- |
|  |  | Stopped using | Still using original | New glasses – free | New glasses – purchased | P-value (univariate) |
|  |  | n=22, 7.9% | n=55, 19.9% | n=85, 30.8% | n=114, 41.3% |  |
| Gender | Female | 1.2 | 10.5 | 74.4 | 14.0 | <0.001 |
|  | Male | 11.1 | 23.8 | 11.1 | 54.0 |  |
| Age groups | 40-49 | 6.1 | 23.5 | 46.9 | 23.5 | <0.001 |
|  | 50-59 | 7.0 | 19.3 | 19.3 | 54.4 |  |
|  | 60-69 | 9.6 | 19.2 | 28.9 | 42.3 |  |
|  | ≥70 | 25.0 | 0.0 | 16.7 | 58.3 |  |
| Personal income | Dependent on other | 5.9 | 10.8 | 64.7 | 18.6 | <0.001 |
|  | Employment or income | 9.2 | 25.3 | 10.9 | 54.6 |  |
| Main use | Household duties | 3.9 | 16.9 | 67.5 | 11.7 | <0.001 |
|  | Phone | 13.9 | 27.8 | 16.7 | 41.7 |  |
|  | Reading | 6.5 | 20.7 | 21.7 | 51.1 |  |
|  | N/A | 80.0 | 20.0 | 0.0 | 0.0 |  |
|  | Working | 0.0 | 18.0 | 11.5 | 70.5 |  |
| Frequency of use | Never/stopped | 100.0 | 0.0 | 0.0 | 0.0 | <0.001 |
|  | Rarely | 18.6 | 44.2 | 9.3 | 27.9 |  |
|  | Few times /week | 2.0 | 17.7 | 60.8 | 19.6 |  |
|  | Daily | 4.8 | 14.4 | 15.2 | **65.6** |  |
| **Knowledge of where to get new glasses** | No | **15.4** | 29.7 | 30.8 | 24.2 | <0.001 |
|  | Yes | 4.3 | 15.1 | 30.8 | **49.7** |  |
| **Reported travel time** | Don't know | 2.9 | 17.6 | **55.9** | 23.5 | <0.001 |
|  | < 30 minutes | 4.3 | 20.0 | 25.7 | **50.0** |  |
|  | 30-60 minutes | 4.9 | 14.8 | **40.7** | **39.5** |  |
|  | 1-2 hours | 0.0 | 8.0 | 4.0 | **88.0** |  |
|  | >2 hours | **25.0** | 12.5 | **37.5** | 25.0 |  |
| **Willingness/ability to pay** | No | 7.3 | 21.5 | 37.0 | 34.3 | <0.001 |
|  | Yes | 4.6 | 18.6 | 33.5 | 43.4 |  |

\*Numbers in the table are row percentages (the sum of percentages in each row is 100%), i.e. distribution of outcome(status of NV glasses) among different demographic/economic groups.

Younger groups had higher proportions still using the original pair and lower discontinuation. Conversely, older adults (≥70) had the highest discontinuation rate (25%) and no participants in this group still used their original glasses, yet they had the highest purchase rate (58.3%).

Individuals with independent income were more likely to purchase their replacements (54.6%) but also had higher rates of discontinuation (9.2%) - mostly due to loss or breakage. Those who were financially dependent mostly received free glasses (64.7%) and had lower discontinuation (5.9%).

Daily users had high replacement rates (65.6% purchased, 15.2% free replacements) and very low discontinuation (4.8%), while those who rarely or never used glasses had minimal replacement and higher discontinuation. Knowing where to get glasses and shorter travel times were associated with replacement. Those unaware of where to access glasses had higher discontinuation (15.4%).

Participants travelling less than 30 minutes to access new glasses showed a 50% replacement purchase rate and low discontinuation (4.3%), compared to those travelling over 2 hours, who had higher discontinuation (25%) and lower replacement (25%) (p < 0.001).

A high proportion of participants (88.9%) expressed willingness and ability to pay for near vision glasses in the future, which was associated with increased rates of purchasing replacements (43.4% among those willing to pay versus 34.3% among those unwilling). Over 94% said they plan on purchasing their next pair.

### Multivariate analysis

#### Factors associated with the continued use of near vision glasses

A logistic regression model was conducted to examine factors associated with the continued use of near vision glasses (table 3). The overall model was statistically significant (LR χ²(12) = 23.14, *p* = 0.02), with a pseudo R² of 0.31, suggesting moderate explanatory power i.e the model explains about 32% of the variation in whether people continue using their glasses.

**Table 3:**
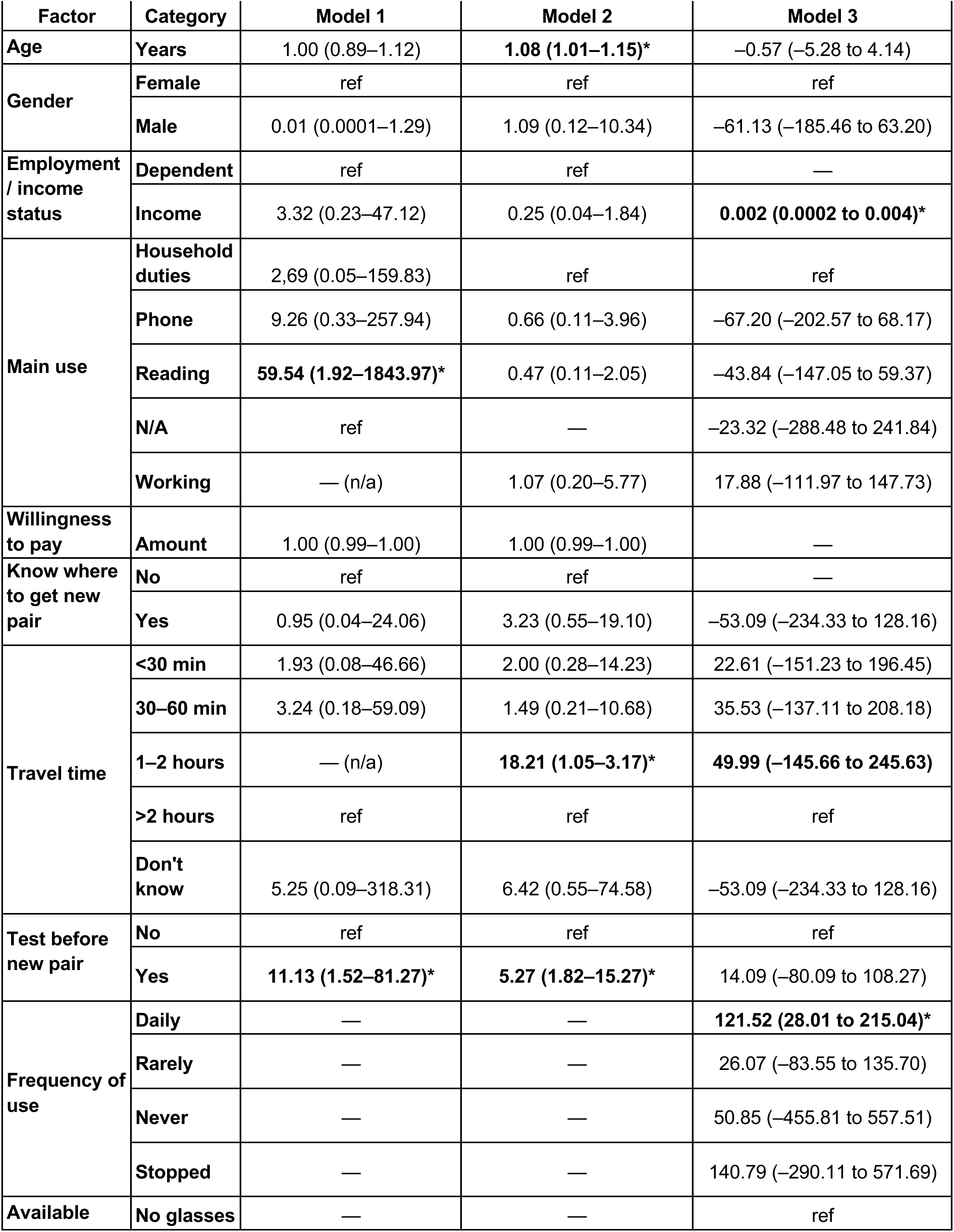
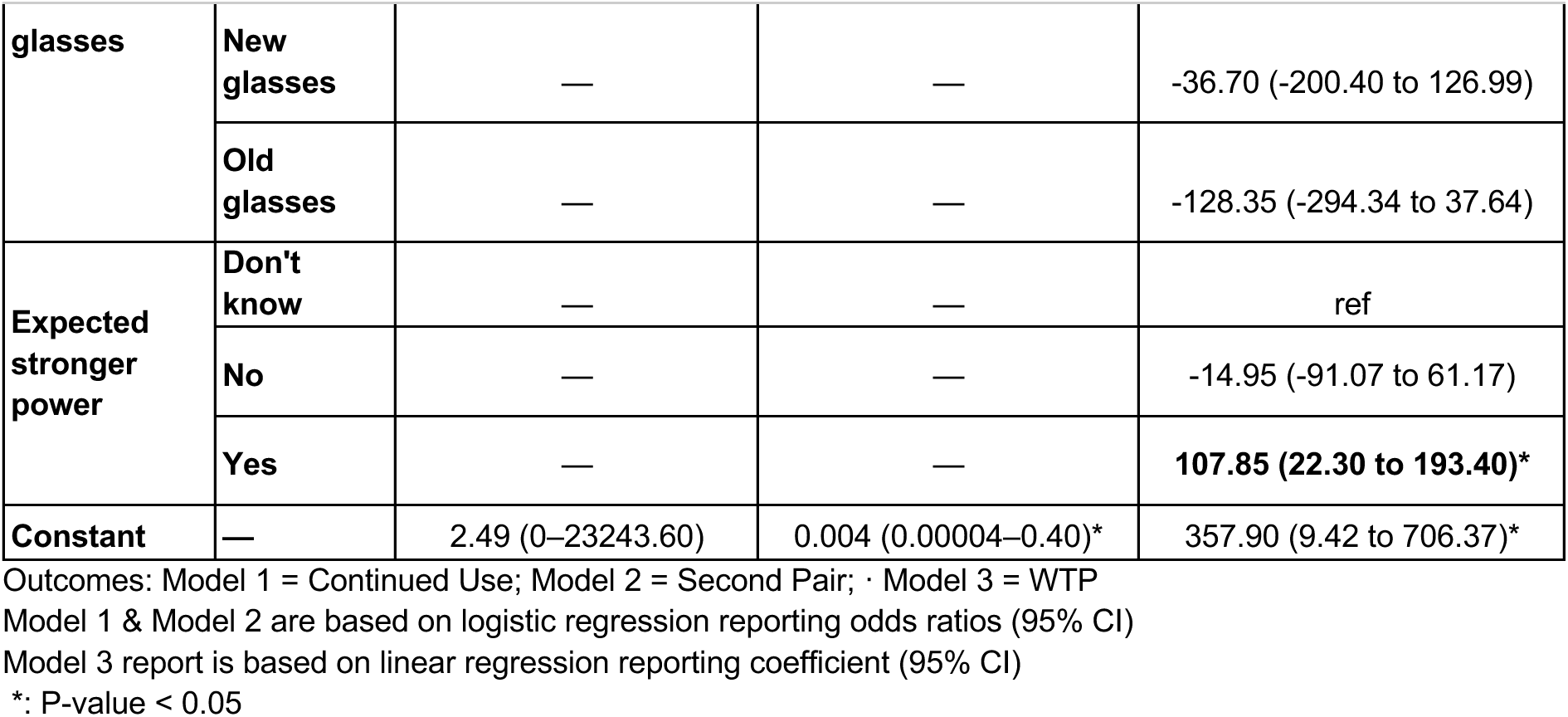
Summary of predictors across three multivariate analysis models.

People continued to use their near vision glasses regardless of age, employment, willingness-to-pay amount, knowledge of where to get new glasses, or travel time to obtain glasses. Gender showed a borderline association: compared to women, men were less likely to continue using glasses (OR = 0.013, 95% CI: 0.0001–1.287, *p* = 0.064). If primarily used for reading, participants were substantially more likely to continue using their glasses (OR = 59.54, 95% CI: 1.92–1843.97, *p* = 0.020). Other uses (household duties, phone) were not statistically significant. Having an eye test before receiving new glasses was strongly associated with continued use (OR = 11.13, 95% CI: 1.52–81.27, *p* = 0.018).

### Factors associated with participants’ willingness-to-pay (WTP) for near-vision glasses

As shown in Table 3, a linear regression model was conducted to assess factors associated with participants’ willingness-to-pay (WTP) for near vision glasses. The model is statistically significant overall. The model explains about 26% of the variation in WTP amount (R-squared = 0.26).

Daily users were more willing to pay compared to occasional users (Coef = 121.52, 95% CI: 28.01–215.04, *p* = 0.011). Income amount was positively associated with WTP: higher income was linked to greater willingness-to-pay (Coef = 0.00197, 95% CI: 0.00016–0.00379, p = 0.033). Expectation of stronger power was significantly associated: those who expected stronger glasses were more willing to pay (Coef = 107.85, 95% CI: 22.30–193.40, p = 0.014). Age, current available glasses, gender, main use of glasses, travel time to get glasses, and having an eye test prior to obtaining new glasses were not significantly associated with WTP.

### Factors associated with obtaining a second pair of near vision glasses

Factors associated with obtaining a second pair of near vision glasses are identified in Table 3. This logistic regression model was statistically significant (LR χ²(13) = 36.57, *p* = 0.0005), with a pseudo R² of 0.178, indicating modest explanatory power.

Age was positively associated with obtaining a second pair: each additional year increased the odds (OR = 1.08, 95% CI: 1.01–1.15, *p* = 0.016). Willingness-to-pay (WTP) showed a borderline positive association (OR = 1.002, *p* = 0.052). Having an eye test before obtaining new glasses was strongly associated with getting a second pair (OR = 5.27, 95% CI: 1.82– 15.27, *p* = 0.002). Travel time showed mixed effects: participants reporting 1–2 hours travel time were significantly more likely to obtain a second pair (OR = 18.21, 95% CI: 1.05–316.95, *p* = 0.047). Other travel-time categories were not significant. Gender (OR = 1.09, *p* = 0.938), employment/income (OR = 0.25, *p* = 0.176), main use of glasses, and knowledge of where to obtain new glasses (OR = 3.23, *p* = 0.195) were not significantly associated with obtaining a second pair.

## Discussion

The vast majority of respondents were still using near vision glasses: a replacement (72.1%), or the original pair (19.9%). Across the regression analyses, several factors emerged as significantly associated with continued use, obtaining a second pair, or willingness-to-pay for near vision glasses. Continued use was more likely among participants who primarily used their glasses for reading and among those who had undergone an eye test before receiving new glasses, highlighting the importance of perceived utility in sustaining use. Obtaining a second pair was positively associated with older age: participants who were older were more likely to replace their glasses, and again strongly linked to having had an eye test. In addition, participants who reported a travel time of one to two hours were unexpectedly more likely to obtain a second pair, though this should be interpreted with caution given the wide confidence intervals. Finally, willingness-to-pay was higher among those with greater income, those using their glasses daily, and those expecting to need a stronger lens power, suggesting that both economic capacity and perceived benefit influence financial investment in replacement glasses. Together, these findings underscore the critical role of functional benefit (reading and daily use) and affordability in promoting sustained and repeat uptake of near-vision correction. Global evidence shows that even modest costs can act as a barrier for the most vulnerable populations, highlighting the importance of affordable pricing or subsidies to ensure equitable access to replacement glasses for all who need them.^10^

Study limitations include, as with all retrospective surveys, potential recall bias regarding the timing, cost and circumstances of replacement. Participants who could not be traced or who declined to participate may differ systematically from those included in the sample, potentially underestimating discontinuation rates. We did not perform clinical reassessment to verify presenting vision status or continued appropriateness of near vision correction, which may limit interpretation of satisfaction and perceived need. Finally, as a cross-sectional study, associations should not be interpreted as causal, and longitudinal follow up would be required to confirm behavioural drivers.

When compared with other similar studies, the findings in Punjab are broadly consistent. In Sierra Leone, 86% of participants were still using near-vision glasses 36–60 months after distribution, either the original (34.9%) or a replacement pair (50.7%).^11^ Most (two-thirds) obtained replacements from hospitals or health centers at modest costs, and more than 95% reported overwhelmingly positive opinions of their glasses. Over half (57.6%) stated that the glasses improved their ability to work or earn income. These results mirror the high retention, replacement behavior, and perceived utility observed in Punjab, reinforcing that affordable access and sustained satisfaction can support long-term uptake across different low-resource settings.

Similarly, evidence from India demonstrated strong and durable demand.^12^ Up to six years after distribution, 93% of participants were still using VisionSpring glasses, 61% had purchased at least one replacement pair, and 88% planned to continue wearing glasses. High satisfaction levels (88%) and reported improvements in near vision (85%) were consistent with findings from Punjab, where willingness-to-pay and perceived benefit were also high. In India, respondents reported an average use for near vision glasses of 4 years, with 32% still using their original pair. In our study also, 31.5% still have/use their original glasses.

Importantly, the Indian data showed that access to a local eye care provider substantially increased the likelihood of purchasing replacements, a finding that aligns with our observation that knowledge of where to obtain glasses was a key determinant of sustained use in Punjab. Furthermore, dissatisfaction or durability issues in India often prompted earlier replacement, highlighting the importance of product quality—consistent with our finding that breakage and loss, rather than dissatisfaction, were the main reasons for discontinuation.

Overall, the Punjab findings add to the growing evidence from South Asia and sub-Saharan Africa that near vision correction programs can achieve high levels of long-term adherence. Longitudinal tracking should be used to monitor usage patterns over time, while survey tools require ongoing refinement to minimize bias. Simple interventions, such as SMS reminders, can encourage continued eyecare, and access-enabling strategies—including vision camps or financial support for optical providers—can further improve uptake.

### Conclusions

This research helps to answer a question often raised by eye care organizations and funders: if we provide near vision glasses for presbyopia, will people continue to use them, and replace them when needed? The study found that among adults aged 40 and over in Punjab, Pakistan, who had received near vision glasses through a community program 3-5 years prior, the vast majority (92.0%) currently owned glasses, with 72.1% having replaced their original pair. 67.0% (95% CI: 61.1-72.5%) knew where to obtain new glasses if needed. A large majority (88.9%; 95% CI: 84.6-92.4%) indicated a positive willingness-to-pay for glasses. Sustained use was high, particularly among those with daily need for use, employment, and knowledge of where to access replacement glasses. Male gender, older age, and personal income were strongly associated with purchasing additional pairs, while women and those dependent on others relied more on free provision. Accessibility, indicated by shorter travel times and awareness of supply sources, and prior eye examinations were also associated with sustained replacement. Furthermore, willingness and ability to pay were high among participants, suggesting good potential for sustained demand beyond initial distribution.

## Declarations

### Ethics approval and consent to participate

The study was approved by COAVS Ethical Review Board [ref: ERB/12A/25], with stakeholder permission to use anonymized patient data. All participants gave informed verbal consent to participate. The study was conducted in accordance with the tenets of the Declaration of Helsinki.

### Consent for publication

Not applicable.

### Availability of data and materials

The datasets generated and analyzed during the current study are not publicly available due to data protection and security agreements, but may be available from the corresponding author on reasonable request.

### Authors’ contributions

All authors contributed to study design and planning. MM, RY, SM and ZHA led running of the study in the field. MK performed the analysis. EW, SLA and MK took the lead in writing the manuscript. All authors discussed the results and provided critical feedback to the final manuscript.

## Acknowledgements

The authors would like to thank all participants, and the staff of COAVS, for their time on this study, as well as the Livelihood Impact Fund for support in the planning and analysis of this work.

## Supplementary Results

**Supplementary Table 1:**
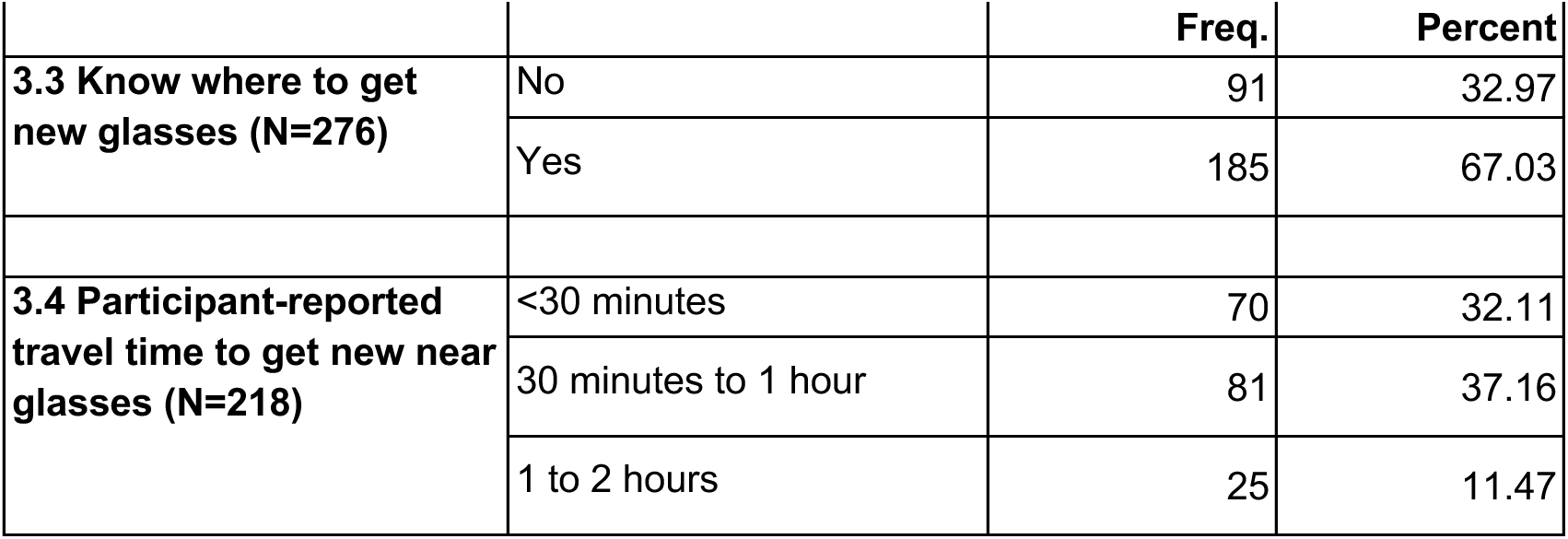
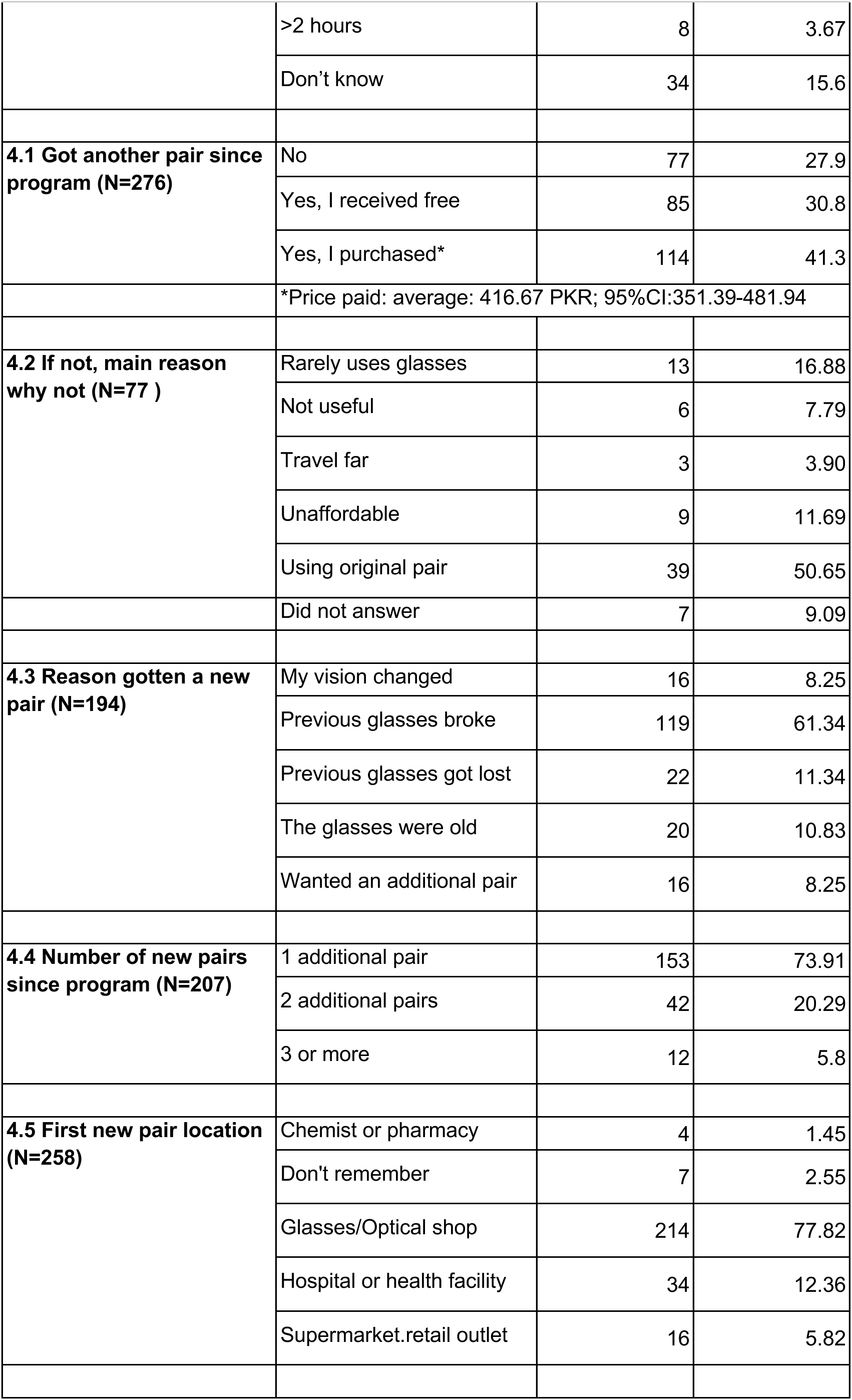
Second pair of glasses after Peek-powered Program.

|  |  |  |  |
| --- | --- | --- | --- |
|  |  | Freq. | Percent |
| 3.3 Know where to get new glasses (N=276) | No | 91 | 32.97 |
|  | Yes | 185 | 67.03 |
| 3.4 Participant-reported travel time to get new near glasses (N=218) | <30 minutes | 70 | 32.11 |
|  | 30 minutes to 1 hour | 81 | 37.16 |
|  | 1 to 2 hours | 25 | 11.47 |
|  | >2 hours | 8 | 3.67 |
|  | Don't know | 34 | 15.6 |
| **4.1 Got another pair since program (N=276)** | No | 77 | 27.9 |
|  | Yes, I received free | 85 | 30.8 |
|  | Yes, I purchased* | 114 | 41.3 |
|  | *Price paid: average: 416.67 PKR; 95%CI:351.39-481.94 |  |  |
| **4.2 If not, main reason why not (N=77 )** | Rarely uses glasses | 13 | 16.88 |
|  | Not useful | 6 | 7.79 |
|  | Travel far | 3 | 3.90 |
|  | Unaffordable | 9 | 11.69 |
|  | Using original pair | 39 | 50.65 |
|  | Did not answer | 7 | 9.09 |
| **4.3 Reason gotten a new pair (N=194)** | My vision changed | 16 | 8.25 |
|  | Previous glasses broke | 119 | 61.34 |
|  | Previous glasses got lost | 22 | 11.34 |
|  | The glasses were old | 20 | 10.83 |
|  | Wanted an additional pair | 16 | 8.25 |
| **4.4 Number of new pairs since program (N=207)** | 1 additional pair | 153 | 73.91 |
|  | 2 additional pairs | 42 | 20.29 |
|  | 3 or more | 12 | 5.8 |
| **4.5 First new pair location (N=258)** | Chemist or pharmacy | 4 | 1.45 |
|  | Don't remember | 7 | 2.55 |
|  | Glasses/Optical shop | 214 | 77.82 |
|  | Hospital or health facility | 34 | 12.36 |
|  | Supermarket.retail outlet | 16 | 5.82 |

**Supplementary Table 2:**
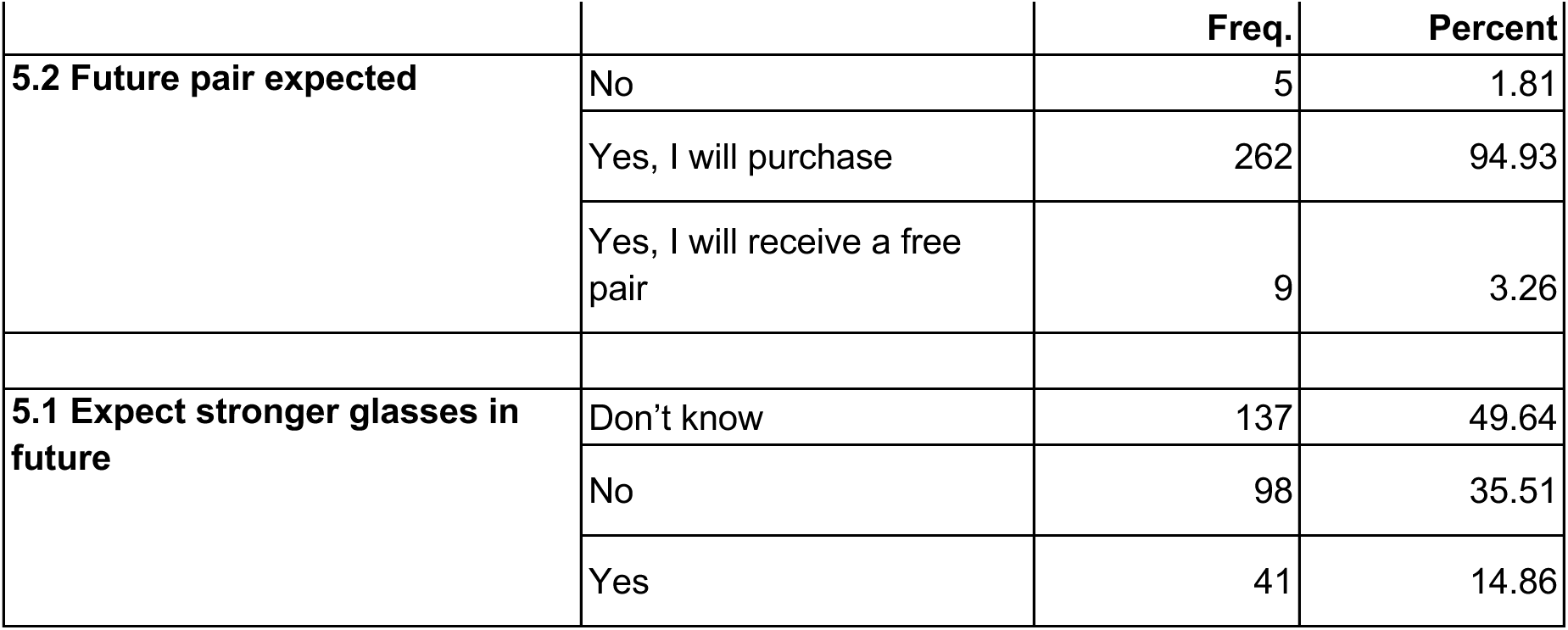
Expectations regarding future NV Glasses (N=276).

**Supplementary Table 3:** Participant-reported willingness to pay for new glasses (Pakistani Rupees).

|  |  |  |  |  |  |  |  |  |  |  |  |
| --- | --- | --- | --- | --- | --- | --- | --- | --- | --- | --- | --- |
| Willingness to Pay* | Respondents (n) | Mean | Min | P.5 | P.25 | P.50 (Median) | P.75 | P.95 | Max | 95% CI |  |
|  |  |  |  |  |  |  |  |  |  | Lower | Upper |
| All participants | 272 | 353 | 0 | 0 | 200 | 300 | 500 | 700 | 1000 | 327 | 379 |
| If WTP is not 0 | 242 | 396 | 100 | 150 | 250 | 350 | 500 | 700 | 1000 | 372 | 421 |

**Supplementary Table 4:** Factors associated with continued use of near-vision glasses after the program.

|  |  |  |  |  |  |  |  |
| --- | --- | --- | --- | --- | --- | --- | --- |
| Factors relation to continued use |  | Odds Ratio | Std. Err. | z | P> \|z\| | 95% CI |  |
| Age | Years | 1.00 | 0.06 | -0.08 | 0.93 | 0.89 | 1.12 |
| Gender | Female | 1.00 | ref |  |  |  |  |
|  | Male | 0.01 | 0.03 | -1.85 | 0.06 | 0.0001 | 1.29 |
| Employment / Income | Dependent on other | 1.00 | ref |  |  |  |  |
|  | Employed or have income | 3.32 | 4.49 | 0.89 | 0.38 | 0.23 | 47.12 |
| Main use | Household duties | 2.69 | 5.61 | 0.47 | 0.64 | 0.05 | 159.83 |
|  | Phone | 9.26 | 157.21 | 1.31 | 0.19 | 0.33 | 257.94 |
|  | Reading | 59.54 | 104.29 | 2.33 | 0.02 | 1.92 | 1843.97 |
|  | N/A | 1.00 | ref |  |  |  |  |
|  | Working | 1.00 | n/a | n/a | n/a | n/a | n/a |
| Willingness to Pay | WTP amount | 1.00 | 0.00 | -1.14 | 0.25 | 0.99 | 1.00 |
| Know where to get new pair | No | 1.00 | ref |  |  |  |  |
|  | Yes | 0.95 | 1.56 | -0.03 | 0.97 | .0372993 | 24.06 |
| Travel time | < 30 min | 1.93 | 3.14 | 0.40 | 0.69 | 0.08 | 46.66 |
|  | 30-60 min | 3.24 | 480.26 | 0.79 | 0.43 | 0.18 | 59.09 |
|  | 1—2 hours | 1.00 | n/a | n/a | n/a | n/a | n/a |
|  | More than two hours | 1.00 | ref |  |  |  |  |
|  | Don't know | 5.25 | 11.00 | 0.79 | 0.43 | 0.09 | 318.31 |
| Test before new pair | No | 1.00 | ref |  |  |  |  |
|  | Yes | 11.13 | 11.29 | 2.38 | 0.02 | 1.52 | 81.27 |
|  | _cons | 2.49 | 11.60 | 0.20 | 0.85 | 0.00 | 23243.60 |

## List of Abbreviations

CI: confidence interval
COAVS: College of Ophthalmology and Allied Vision Sciences
CRET: Competency-based Refractive Error Teams
NVI: near vision impairment
OR: odds ratio
PKR: Pakistani rupee
WTP: willingness-to-pay
USD: United States dollar

